# Retinal Transcriptome-Wide Association Study Identifies Novel Alzheimer’s Disease Risk Genes

**DOI:** 10.64898/2026.07.01.26357036

**Authors:** Yinuo Zhang, Brian Chen, Huaigu Sun, Ruhao Shi, Yun Li, Jiawen Du, Andrea R.V.R. Horimoto

**Affiliations:** Department of Biostatistics, University of North Carolina at Chapel Hill, Chapel Hill, NC, 27599, USA; School of Public Health, Rutgers University, Piscataway, NJ, 08854, USA; Department of Genetics, University of North Carolina at Chapel Hill, Chapel Hill, NC, 27599, USA

**Keywords:** Alzheimer’s disease, transcriptome-wide association study (TWAS), retina, expression quantitative trait loci (eQTL), complement system, OTTERS

## Abstract

**INTRODUCTION:** Alzheimer’s disease (AD) is the leading cause of dementia worldwide. The retina shares molecular pathways with the brain, yet no study has systematically linked retinal gene expression to AD risk.

**METHODS:** We performed transcriptome-wide association studies (TWAS) using two independent retinal eQTL panels (Strunz et al., n = 311; EyeGEx, n = 406) and a large meta-analyzed AD genome-wide association study (GWAS) (Bellenguez et al., 111,326 cases, 677,663 controls). Genes were further validated with GWAS in the independent Alzheimer’s Disease Sequencing Project (ADSP) using a matched eQTL-panel strategy.

**RESULTS:** We identified 62 AD-associated genes across the two eQTL panels using Bellenguez et al. as the discovery cohort. Of these, 31 were replicated in the ADSP cohort. The findings highlight shared complement-mediated immune dysregulation (*CD55*, *CD46, TREM2*) and provide functional transcriptomic evidence to prioritize novel causal drivers of AD pathogenesis, including *STYX* and the *LRRC37* gene family.

**DISCUSSION:** Retinal data capture core AD genetic architecture and reveal novel risk genes, highlighting the retina as a molecularly informative tissue for dementia research.

## Background

Alzheimer’s disease (AD) is the most common form of dementia, affecting over 55 million individuals worldwide, with prevalence projected to exceed 150 million by 2050 [1]. Although traditionally viewed as a disorder strictly of the brain, a growing body of evidence shows that AD pathology also appears in peripheral tissues, specifically the retina [2,3]. As a developmental outgrowth of the diencephalon, the retina shares embryological origins, anatomical features, and molecular pathways with the central nervous system (CNS). Retinal abnormalities documented in AD patients include thinning of the retinal nerve fiber layer, vascular changes, and amyloid-beta deposition [3–5]. Optical coherence tomography (OCT) measures of retinal structure have been proposed as potential noninvasive biomarkers for AD screening [6].

While current genome-wide association studies (GWAS) have identified over 75 genetic loci associated with AD risk, translating these findings into clear biological insights remains challenging7/1/2026 11:32:00 AM[7,8]. Transcriptome-wide association studies (TWAS) offer a powerful framework to bridge this gap by integrating GWAS with expression quantitative trait loci (eQTL) data to identify genes whose genetically regulated expression associates with disease risk [9,10]. Prior TWAS for AD have predominantly employed brain tissue eQTL references, including cortical, hippocampal, and cell-type-specific panels, and have successfully implicated genes at established AD loci as well as novel candidates [11,12]. However, brain tissue eQTL panels are constrained by limited sample sizes, challenges in postmortem collection, and limited cell-type resolution, motivating the investigation of complementary tissue contexts that may capture additional disease-relevant gene regulatory variation [13].

The retina presents a compelling alternative tissue for AD TWAS [2]. Beyond its shared developmental origin with the brain, the retina harbors microglia-like resident macrophages, expresses many of the same synaptic and immune-related genes implicated in AD, and is the only CNS-derived tissue accessible to noninvasive imaging in living patients [2,5,14]. Specifically, among the shared immune mechanisms, complement pathway dysregulation is a hallmark of both AD and age-related macular degeneration (AMD), and provides a mechanistic link between retinal and brain neurodegeneration [7,15,16]. Recent advances in retinal genomics have produced comprehensive eQTL datasets from retinal tissue [17,18], enabling tissue-specific TWAS analyses that may uncover AD-associated gene regulation not captured by brain-focused resources.

Here, we conducted a retinal TWAS for AD using two independent retinal eQTL reference panels: a mega-analysis retinal eQTL dataset derived from Strunz et al. [18] (hereafter Meta-eQTL) and the Eye Genotype Expression (EyeGEx) dataset [17]. Each eQTL panel was paired separately with the largest available AD GWAS meta-analysis [8] (hereafter Bellenguez et al.) using the Omnibus Transcriptome Test using Expression Reference Summary data (OTTERS) [19] framework to evaluate whether genetically predicted retinal gene expression influences AD susceptibility. We also performed independent validation in the Alzheimer’s Disease Sequencing Project (ADSP) cohort using a matched eQTL-panel strategy [20]. Our analysis validated 31 genes whose genetically regulated retinal expression is associated with AD, including well-established and novel candidates, which have not been previously reported in GWAS or brain-tissue TWAS for AD. Together, these transcriptomic signatures provide evidence for complement-mediated neurodegeneration as a convergent retina-brain AD signal.

## Methods

### AD GWAS summary statistics

For discovery, we used the largest AD GWAS meta-analysis in individuals of European ancestry to date [8] (Bellenguez et al; 111,326 cases and 677,663 controls, ∼21M variants), which comprised both clinically diagnosed cases and proxy cases (i.e., individuals with parental history of AD or dementia).

For validation, we conducted a GWAS in the ADSP cohort (6,730 clinically diagnosed cases and 5,381 controls of self-reported White). We restricted our analysis to variants with minor allele frequency (MAF) > 0.01 and genotype missingness < 0.05. For 991 individuals older than 90, we capped their age at 90, and for individuals with missing age data, we imputed with the mean age (73.9 years). We then conducted a GWAS for AD using REGENIE [21], adjusting for age, sex, and the top 10 principal components (PCs).

### Retinal eQTL panels

We used two independent retinal *cis*-eQTL reference panels: (1) Meta-eQTL, a mega-analysis of 311 healthy postmortem retinas of European ancestry across three sites conducted by Strunz et al [18]; and (2) the EyeGEx panel [17] comprising 406 postmortem retinas spanning controls (MGS1) and age-related macular degeneration (AMD) stages MGS2-4. Both panels were derived from bulk retinal RNA-sequencing without cell-type deconvolution, with *cis*-eQTL windows defined as ±1 Mb of the gene transcription start site (TSS), and underwent QC by their original studies prior to public release. Only autosomal genes with available *cis*-eQTL summary statistics were retained for TWAS. Detailed panel characteristics, gene-level QC, and Ensembl GRCh38 gene-annotation procedures are reported in the **Supplementary Methods**.

### TWAS analysis using OTTERS

We performed TWAS using the OTTERS [19] in a two-stage framework. Stage 1 leverages retinal eQTL summary statistics to train multiple expression-prediction models, and stage 2 integrates these models with AD GWAS data to evaluate gene expression-disease associations in FUSION Z-score, and aggregate results into a single statistical inference via an Aggregated Cauchy Association Test (ACAT-O) [22] (details see **Supplementary Methods**). Separate TWAS analyses were conducted for each combination of GWAS datasets (Bellenguez et al., ADSP) and eQTL panels (Meta-eQTL, EyeGEx).

To account for multiple testing, we applied Bonferroni corrections based on the number of genes successfully tested in each eQTL panel. Specifically, we tested 16,559 genes in the Meta-eQTL panel and 17,071 genes in the EyeGEx, yielding Bonferroni-corrected significance thresholds of 3.02 × 10^-6^ and 2.93 × 10^-6^, respectively (**Supplementary Table 1**).

### Gene validation and prioritization

AD-associated genes were defined as those achieving Bonferroni significance in Bellenguez et al. + Meta-eQTL or Bellenguez et al. + EyeGEx (or both). To control for eQTL model-specific noise, we applied a matched-panel validation strategy: discovery genes were considered validated at a lenient threshold, exceeding nominal significance (p < 0.05) in the ADSP cohort using the same retinal eQTL panel, as well as having a same effect direction (details see below). A gene was considered novel if it had not been previously reported in AD GWAS nor in prior brain-tissue TWAS of AD.

### Cross-cohort directional concordance analysis

To ensure the validity of our validation, we assessed the consistency of the effect direction between the discovery and validation cohorts, by evaluating the sign of the mean FUSION Z-score, Z□ (defined as the numerator 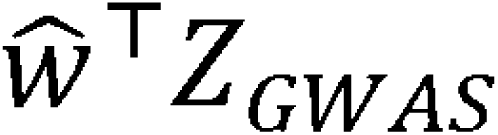 of FUSION Z-score formula, averaged across all five models). For genes with discordant Z□ signs, we first capped the ADSP Z-score at |Z| = 9.3 to match the Z-score cap used in Bellenguez et al., and quantified the fractional contribution of variants within ±700Kb of the *APOE* index variant rs4420638 (window justification in **Supplementary Methods; Supplementary Table 7**). Second, we accounted for differential power between ADSP (N ∼ 12K) and Bellenguez et al. (N ∼ 789K) (full procedure in **Supplementary Methods**). Finally, we consider the unsolved discordance is likely driven by uncaptured LD architecture or cohort-specific technical confounding, if the sign reversal persisted after Z-score capping and the residual deviated from zero by more than two standard errors. Otherwise, the directional flip was attributed to technical artifacts such as extreme-value distortion or differential statistical power.

### Pairwise correlations of predicted gene expression, conditional, colocalization and MR analyses

To assess the extent to which TWAS signals at multi-gene loci could be driven by shared predicted expression rather than independent regulatory effects, we computed pairwise Pearson correlations of genetically regulated expression (GReX) between all genes within each locus of interest, using individual-level in the 1000 Genomes [23] European-ancestry reference panel, by multiplying the eQTL weights from the best-performing OTTERS model by individual-level genotype dosages at all model SNPs, then summing across SNPs (**Supplementary Methods**).

Beyond correlation, to further estimate if the genes are clustered or partially independent with each other, we performed forward-selection conditional TWAS at five multi-gene loci (chr1q32 *CD55/CD46*, chr16p11.2 *SEZ6L2/DOC2A*, chr17q21.31 *MAPT/LRRC37A*, chr17q23 *ACE*, and chr19q13 *APOE*; **Supplementary Methods**).

To test whether the retinal eQTL and AD GWAS signals at each locus share a causal variant or are driven by distinct variants in LD, we performed colocalization analysis for all significant genes in either the discovery or validation phase. We used coloc.abf function from R package *coloc* (v5.2.3) [24]. We set a ±1 Mb window around the gene body and extracted eQTL and GWAS summary statistics for all variants in that region. We considered PP.H4 > 0.75 as evidence for a shared causal variant.

Finally, to evaluate causal evidence for novel candidate genes, we performed Mendelian randomization (MR) using eQTL variants associated with the target gene at FDR < 0.1 as instruments. The causal effect was primarily estimated using the random-effects inverse-variance weighted (IVW) method. To assess the robustness of our results against potential violations of MR assumptions, particularly horizontal pleiotropy, sensitivity analyses were performed using MR-Egger regression [25].

## Results

### Study Overview

In this study, we combined summary statistics from the largest AD GWAS meta-analysis to date (Bellenguez et al.) [8] with two independent retinal eQTL datasets, Meta-eQTL (n = 311) and EyeGEx (n = 453), using the OTTERS pipeline (**Figure 1, Supplementary Table 1**). Our goal was to identify genes whose expression in the retina is regulated by genetic variants and is associated with AD risk. We validated our findings in the ADSP cohort. Genes were considered validated if they showed nominal significance (p < 0.05) using the matched eQTL panel.

**Figure 1.**
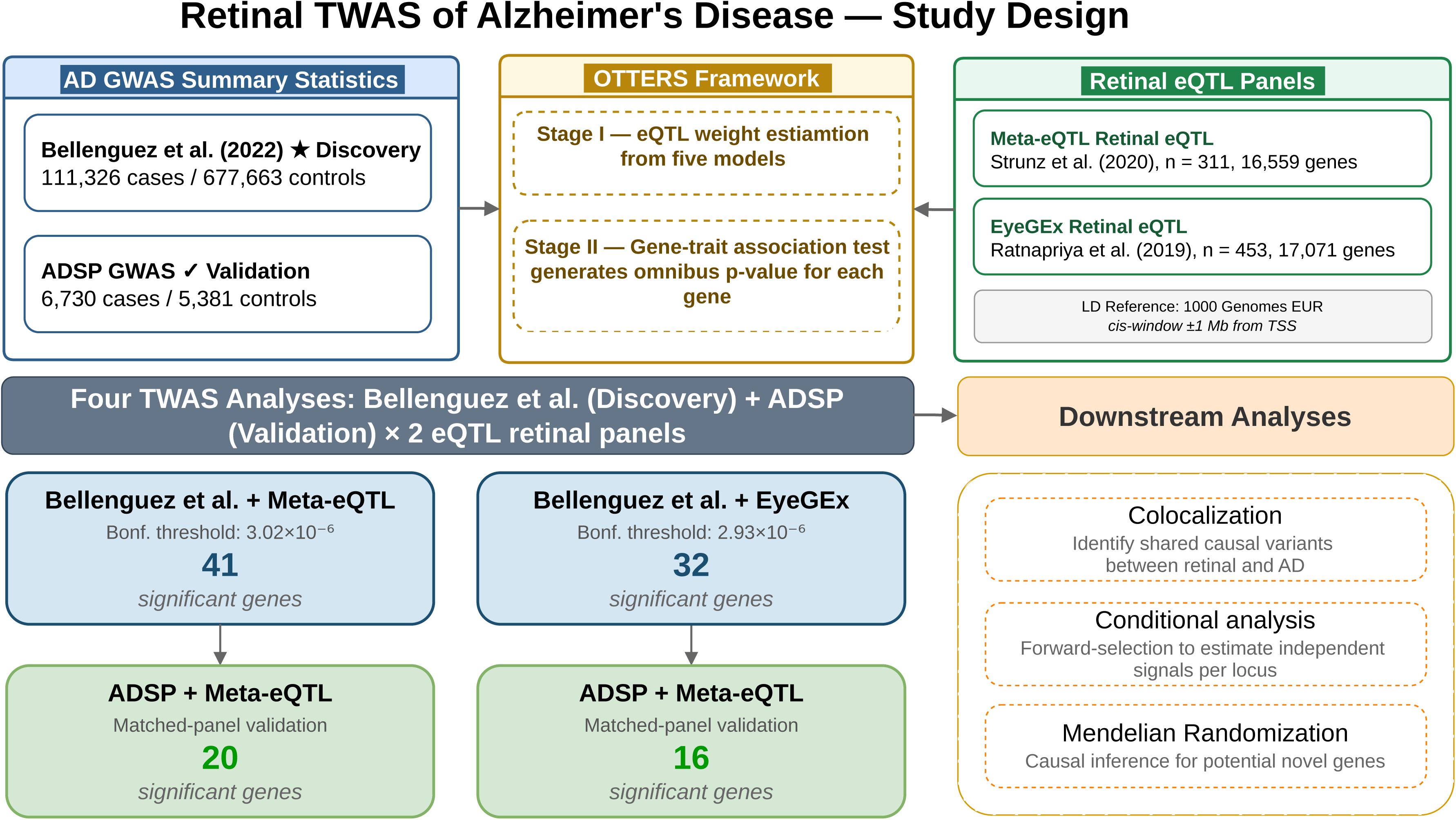
Study design overview. The Bellenguez et al. (2022) GWAS was used as the discovery dataset and integrated with two independent retinal eQTL panels (Meta-eQTL and EyeGEx) using the OTTERS framework. Independent validation was performed in the ADSP GWAS using a matched-eQTL-panel. Numbers of significant findings in discovery and validation analyses are shown. The four genes achieving Bonferroni significance in 3 of 4 analyses are listed as the highest-confidence findings.

### Discovery of AD-associated genes through retinal TWAS

Two discovery analyses, Bellenguez et al. + Meta-eQTL (BM) and Bellenguez et al. + EyeGEx (BE), were conducted. We observed association signals from clear tail enrichment without systematic inflation (λ = 0.746 - 0.768, **Supplementary Figure 1**). BM and BE identified 41 and 32 Bonferroni-significant genes, respectively, yielding 62 unique AD-associated genes in total (**Figure 2A-B, Supplementary Table 2**). Of these, 12 (19%) achieved Bonferroni significance in both panels, 29 were identified only in BM, and 21 only in BE.

**Figure 2.**
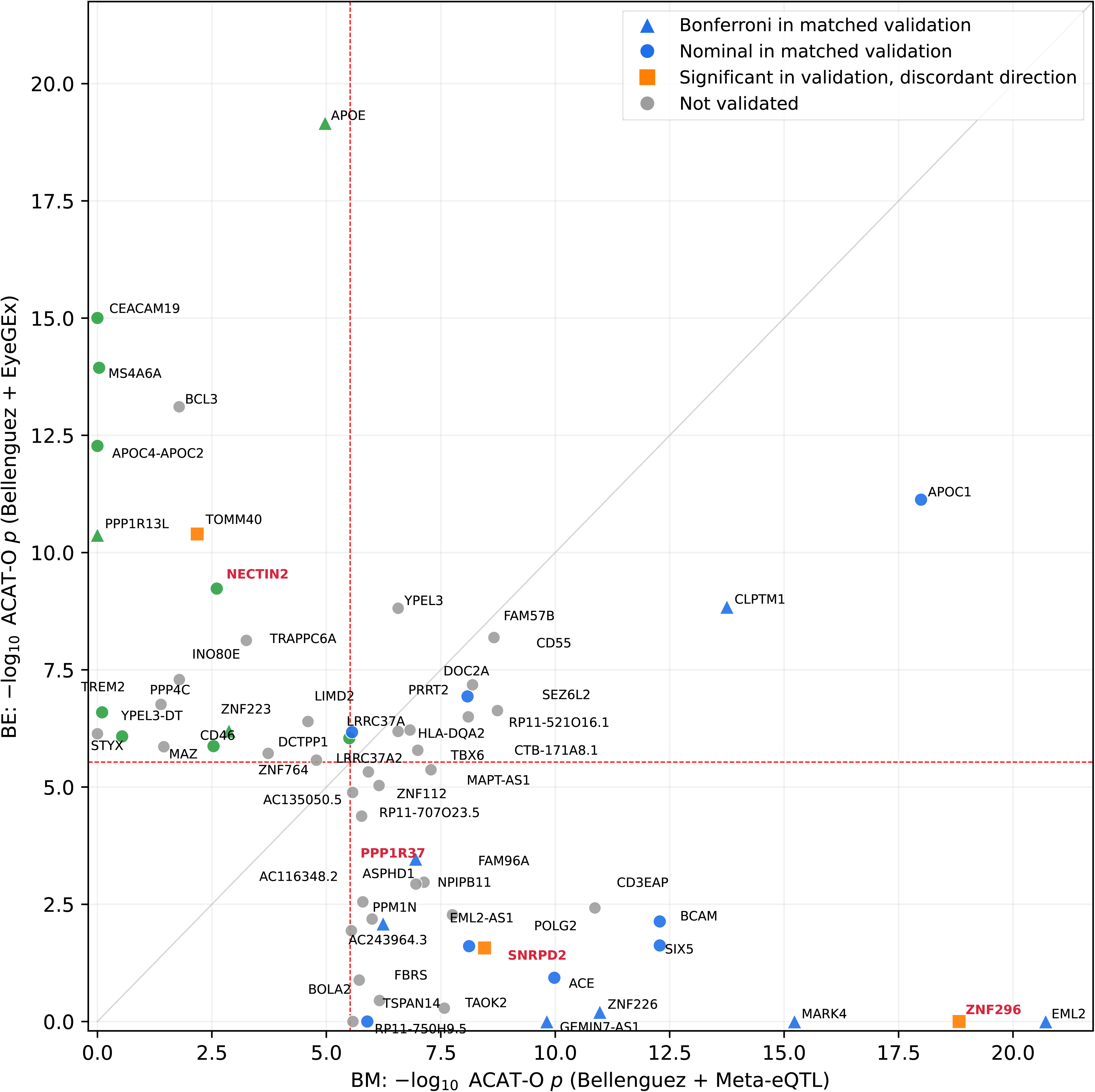
Transcriptome-wide Manhattan plots. Each point represents a gene, plotted by chromosomal position (x-axis) against-log_lO_ of OTTERS’ ACAT-O omnibus p-value (y-axis). Dashed horizontal lines indicate the Bonferroni-corrected significance thresholds (Meta-eQTL panel: 3.02 x 10^-6^; EyeGEx panel: 2.93 x 10^-6^). Genes shown in red bold (SN*RPD2, PPP1R37, ZNF296, NECTIN2*) denote the conditionally independent signals identified at the chromosome 19q13 locus (see Figure 5A **and Supplementary Table 9**).

The 12 genes significant in both panels spanned five chromosomes (**Supplementary Table 2**). Several are well-established AD risk genes, including *APOC1* (p_BM_ = 1.0 × 10^-18^; p_BE_ = 7.4 × 10^-12^) and *CLPTM1* (p_BM_ = 1.8 × 10^-14^; p_BE_ = 1.5 × 10^-9^). *CD55* (p_BM_ = 8.2 × 10^-9^; p_BE_ = 1.2 × 10^-7^), a complement regulatory gene implicated in AMD but not previously reported in AD brain TWAS, was also among the cross-panel discoveries [26,27]. Five of the 12 shared genes mapped to the chr16p11.2 locus (*SEZ6L2*, *DOC2A*, *YPEL3*, *PRRT2*, and *DOC2A*), a region with established neurodevelopmental associations [28].Cross-panel consistent significance for these genes suggests that their regulatory signals are robust to differences in eQTL reference composition.

Among the 29 genes identified exclusively in BM, the strongest signals (*EML2, ZNF296, MARK4, and ZNF226*) clustered on chromosome 19 **(Figure 3A)**. Beyond this cluster, the BM panel also detected the established *ACE* locus and the mitochondrial subunit *POLG2* on chromosome 17, along with *TSPAN14* on chromosome 10, and a novel lncRNA (ENSG00000260634) on chromosome 2.

**Figure 3.**
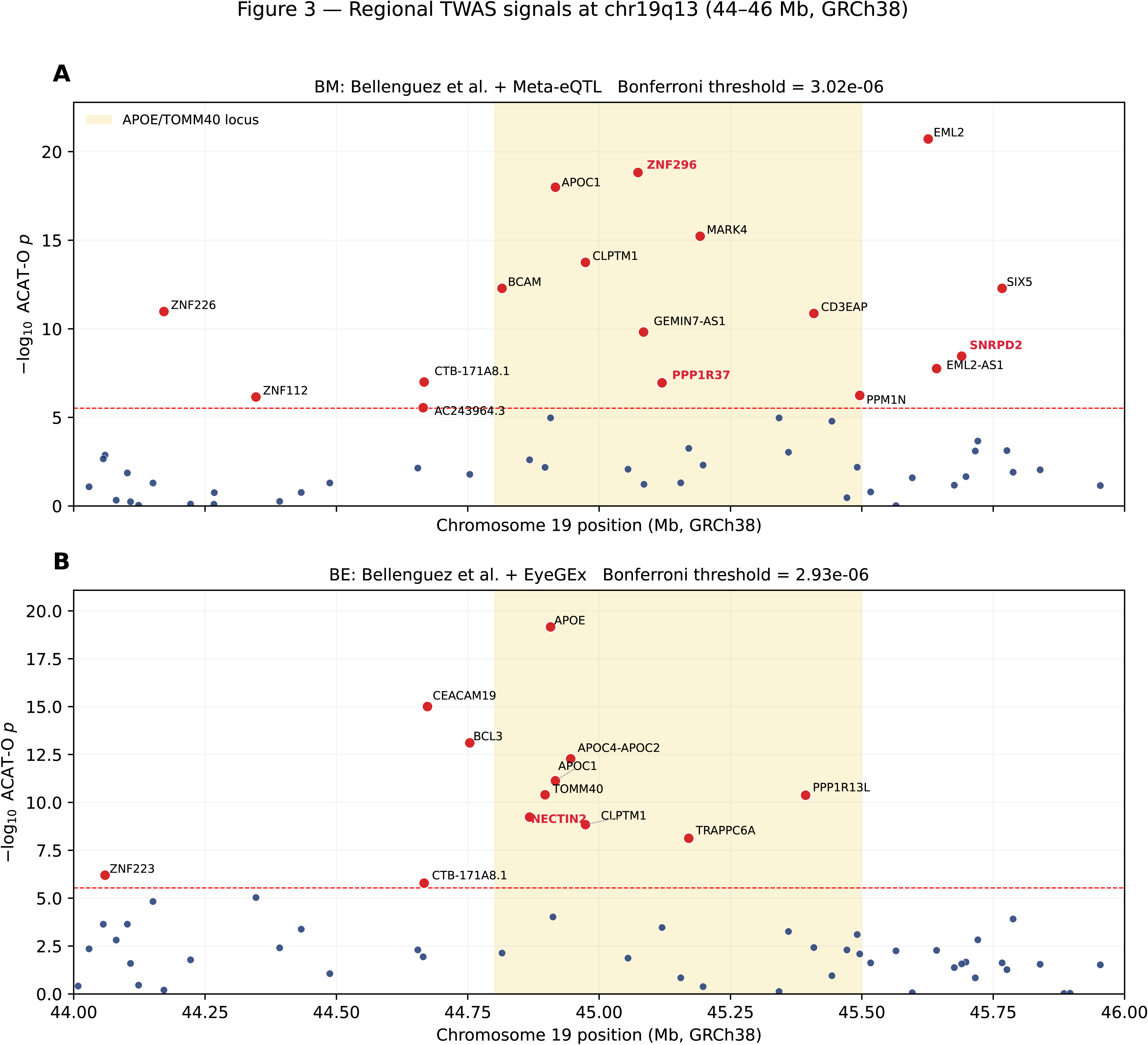
Regional TWAS association signals at the chromosome 19q13 locus (44–55 Mb, GRCh38). Panels show-log_lO_(p) for OTTERS’ ACAT-O gene-level associations from Bellenguez et al. + Meta-eQL (A) and Bellenguez et al. + EyeGEx (B). The dashed horizontal line denotes the Bonferroni-corrected threshold (Meta-eQTL: 3.02 x 10^-6^; EyeGEx: 2.93 x 10^-6^). The shaded region highlights the *APOE/TOMM40* locus. Genes exceeding the Bonferroni threshold in at least one analysis are labeled. Genes shown in red bold (*SNRPD2, PPP1R37, ZNF296, NECTIN2*) denote the conditionally independent signals identified at this locus (see Figure 5A **and Supplementary Table 9**).

The BE analysis contributed 21 unique discoveries. Interestingly, this panel captured several canonical Alzheimer’s genes that did not reach significance in the BM panel, such as *APOE, TOMM40, NECTIN2* **(Figure 3B)**. Outside the chromosome 19 region, BE also uniquely identified the microglial markers *TREM2* and *MS4A6A,* the complement regulator *CD46* (adjacent to *CD55*) and *STYX*, a pseudophosphatase involved in neuronal growth [29,30].

As expected, the majority of signals concentrated on chromosome 19q13, aligning with the known *APOE* risk architecture. Moreover, the presence of two complement regulators, *CD55* and *CD46* on chromosome 1q32, provides convergent evidence for complement-mediated mechanisms in the retina. Although only 12 of the 62 discovery genes reached Bonferroni significance in both panels, panel-exclusive genes were strongly enriched for nominal significance in the other panel: 82% of BE-specific genes (14/17 tested) reached p < 0.05 in BM, and 78% of BM-specific genes (18/23 tested) reached p < 0.05 in BE, compared to 7% in the background (i.e., genes outside the 62-gene discovery set). This pattern indicates that panel non-overlap does not reflect fundamentally distinct biology, and supports our matched same-panel validation strategy.

### Matched-panel validation

For validation, we applied a similar matched-panel strategy, yielding two validation analyses: ADSP + Meta-eQTL (AM) and ADSP + EyeGEx (AE) (**Figure 2C-D**). Among the 12 cross-panel discovered genes, five were successfully validated (*APOC1, CLPTM1, CD55, LRRC37A*, and *CEACAM16-AS1*) (**Supplementary Table 2**). Specifically, *APOC1* is a well-established AD gene confirmed across brain TWAS, blood TWAS, and GWAS [8,12,31]. *LRRC37A* has prior support from brain TWAS through its paralog *LRRC37A2* and from neuroimaging TWAS [32]; our retinal signal extends this locus to a third tissue. While *CD55* has not been reported in prior AD brain TWAS, it encodes a key complement regulator (decay-accelerating factor). Complement dysregulation is implicated in both AMD (where *CD55* has been identified by TWAS) and AD (through microglial C1q/C3-mediated synaptic pruning), and *CD55* has separately appeared in an AD myeloid genomics study [32,33]. The cross-panel retinal signal therefore represents a novel AD finding that aligns with this established complement biology.

For panel-specific findings, 20 of 41 BM discovery genes were validated in AM, and 16 of 32 BE discoveries validated in AE (**Supplementary Table 3**). These rates are consistent with the substantially smaller ADSP sample size compared to the Bellenguez et al. GWAS (∼12K vs. 789K participants). Although our validation criteria required only nominal significance, 7 BM and 4 BE genes further exceeded the more stringent Bonferroni threshold in the validation cohort. The final validated set consisted of 36 discovery-validation pairs, representing 31 unique genes (**Supplementary Tables 4 and 5**).

Four genes mapping to chromosome 19q13 achieved Bonferroni significance across three of four discovery and validation analyses (**Supplementary Table 6**): *CLPTM1*, *TOMM40*, *ZNF226*, and *CEACAM16-AS1*. Among these four genes, only *TOMM40* exhibited a discordant direction of effect (**Supplementary Table 7**), a reversal that likely reflects the extreme LD complexity at the adjacent *APOE* locus as detailed in following EyeGEx panel-specific section. Specifically, *CLPTM1* and *TOMM40* were established AD GWAS and brain TWAS genes [34,35]. *ZNF226* was previously identified in blood-based AD TWAS [31], but our results provide the first tissue-specific evidence for its role in the retina. *CEACAM16-AS1* is a long noncoding antisense RNA transcribed opposite to *CEACAM16*, *CEACAM19*, and *PVR* on chromosome 19q13. While the 19q13 *CEACAM* cluster has been implicated in AD through *CEACAM19* in brain and spleen tissues, our results extend signal within this locus to *CEACAM16-AS1* in retinal tissue [12].

### Meta-eQTL panel-specific validated genes

Of the 29 genes discovered exclusively through the Meta-eQTL panel, 14 were validated in ADSP, of which 10 (71%) have been previously reported in AD GWAS or brain-tissue TWAS [8,12,36]. This proportion is comparable to the EyeGEx-specific validated set (see below), indicating that the Meta-eQTL panel captures a representative cross-section of known AD biology. All validated genes showed consistent direction of effect between discovery and validation, with two exceptions: *ZNF296* and *SNRPD2.* However, our sensitivity analyses confirmed that neither represented a genuine biological reversal. The apparent flip for *ZNF296* was shown to be driven by LD leakage from *APOE*: 45.1% of the *ZNF296* TWAS signal in ADSP came from variants in LD with the *APOE* variant rs4420638, instead of from variants that regulate *ZNF296* itself. Applying the matched Z-score capping reduced its validation signal to null (**Supplementary Table 7**). Similarly, for *SNRPD2*, after applying capping, the flip fell within sample-size-scaled sampling variation after capping (capped validation Z□ = +1.30; **Supplementary Table 7**).

Among the validated genes, we found some novel candidates without prior AD evidence. *POLR1G* (p_BM_ = 1.36 × 10^-11^; p_AM_ = 5.42 × 10^-4^) encodes a subunit of RNA Polymerase I involved in ribosomal biogenesis, a process increasingly linked to neurodegeneration through nucleolar stress [37]. *POLG2* (p_BM_ = 7.61 × 10^-9^; p_AM_ = 3.67 × 10^-3^) encodes the accessory subunit of mitochondrial DNA polymerase γ, which is required for mtDNA replication and maintenance [38]. Disruption of mtDNA maintenance is among the earliest molecular features of AD pathogenesis [39], suggesting a plausible mechanism by which retinal *POLG2* dysregulation could contribute to AD risk. Among noncoding transcripts, *GEMIN7-AS1* (p_BM_ = 1.52 × 10^-10^; p_AM_ = 5.34 × 10^-10^) achieved significance in both discovery and validation even after adjusting for multiple testing. GEMIN7-AS1 is a long noncoding antisense RNA transcribed opposite to the protein-coding *GEMIN7* gene, which was nominally associated with AD in a recent multi-tissue TWAS [40], raising the possibility that antisense-mediated regulation of GEMIN7 expression is the functional mechanism at this locus. ENSG00000279095 (p_BM_ = 2.84 × 10^-6^; p_AM_ = 4.76 × 10^-2^), an unannotated lncRNA on chromosome 19, had no prior AD evidence and represents an entirely novel finding. To further explore the causality for these novel candidates, we conducted MR analysis and found that all of them show a causal relationship with AD, in at least one of four configurations (i.e., BM, BE, AM, AE; **Supplementary Table 8**).

### EyeGEx panel-specific validated genes

Of the 21 genes discovered exclusively through the EyeGEx panel, 11 were validated in ADSP, of which 10 (91%) have prior AD literature support [8,12,36]. Five EyeGEx-specific genes were absent from the Meta-eQTL catalog (*APOE, CEACAM19, APOC4-APOC2, PPP1R13L, YPEL3-DT*), while others, such as *TREM2* (p_BM_ = 0.79) and *MS4A6A* (p_BM_ = 0.92), showed no signal, indicating that the EyeGEx eQTL weights captured regulatory variation at these loci that the Meta-eQTL panel did not. All validated genes showed consistent direction of effect (**Supplementary Table 7**), with the exception of *TOMM40,* which exhibited a sign reversal of effect between discovery and validation. To determine whether this reflected genuine biological discordance, we applied the cross-cohort sensitivity analyses (see **Methods**) and the sign reversal persisted.

The EyeGEx panel recovered several established AD loci not detected by the Meta-eQTL panel, including *MS4A6A, NECTIN2,* and *TREM2*. The identification of *TREM2* in retinal tissue is consistent with the presence of resident myeloid cells in the retina that share activation patterns with brain microglia [41]. The principal finding not previously reported in AD GWAS or brain-tissue TWAS was *STYX* (p_BE_ = 8.32 × 10^-7^; p_AE_ = 4.77 × 10^-2^) on chromosome 14. *STYX* encodes a catalytically inactive pseudophosphatase that regulates neuronal growth and has been implicated in neurodegenerative processes [42]. A recent multi-ancestry AD TWAS identified this locus, though *STYX* was not prioritized as a top gene after fine-mapping [42]. Again, to further investigate the potential causal relationship between *STYX* expression and AD, we performed MR analysis. Our results robustly corroborated the TWAS signal, demonstrating that increased *STYX* expression is associated with elevated AD risk in both Bellenguez et al. (IVW OR = 1.17, 95% CI: 1.15–1.19, p = 6.8 × 10^-65^) and ADSP (IVW OR = 1.12, 95% CI: 1.10–1.15, p = 3.7 × 10^-23^; **Supplementary Table 8**). In conclusion, these findings provide independent tissue-specific evidence for *STYX* and suggest it may contribute to AD pathology through regulatory mechanisms operating in retinal neurons.

### Comparison of eQTL panels and discovery complementarity

Direct comparison of the two discovery analyses reveals largely complementary discovery profiles (**Figure 4**). 12 of the 62 unique discovery genes (19%) reached Bonferroni significance in both BM and BE. Among genes Bonferroni-significant in only one panel, most still reached nominal significance in the other panel: 82% of BE-specific genes (14/17 tested) reached p < 0.05 in BM, and 78% of BM-specific genes (18/23 tested) reached p < 0.05 in BE, compared to 7% in the background gene set, indicating that panel non-overlap does not reflect fundamentally distinct biology. The Meta-eQTL panel yielded stronger associations at several chromosome 19 loci, including *ZNF226* (p_BM_ = 1.1×10^-11^; p_BE_ = 0.63) and *SNRPD2* (p_BM_ = 3.5×10^-9^; p_BE_ = 0.027), indicating that the regulatory variants driving these associations are better represented in the Meta-eQTL reference. The EyeGEx panel, in turn, uniquely recovered several established AD genes that did not reach Bonferroni significance in Meta-eQTL, including *APOE*, *TOMM40*, *MS4A6A*, and *NECTIN2*. One possible explanation is that EyeGEx includes AMD-affected retinas alongside controls, which may expose regulatory variation at immune and lipid metabolism loci that remains inactive in the healthy tissue profiled by the Meta-eQTL panel. Of the 39 discovery–validation pairs reaching nominal significance, 36 showed consistent direction of effect between discovery and validation; the remaining three (*ZNF296*: p_BM_ = 8.0×10^-19^, p_AM_ = 0.026; *SNRPD2*: p_BM_ = 3.5×10^-9^, p_AM_ = 0.027; *TOMM40*: p_BE_ = 4.4×10^-11^, p_AE_ = 5.0×10^-11^) were directionally discordant and excluded from the validated set. Genes reaching Bonferroni significance in both panels, particularly *CD55* and *CEACAM16-AS1*, showed consistent effect sizes across the two references, supporting the robustness of these associations.

**Figure 4.**
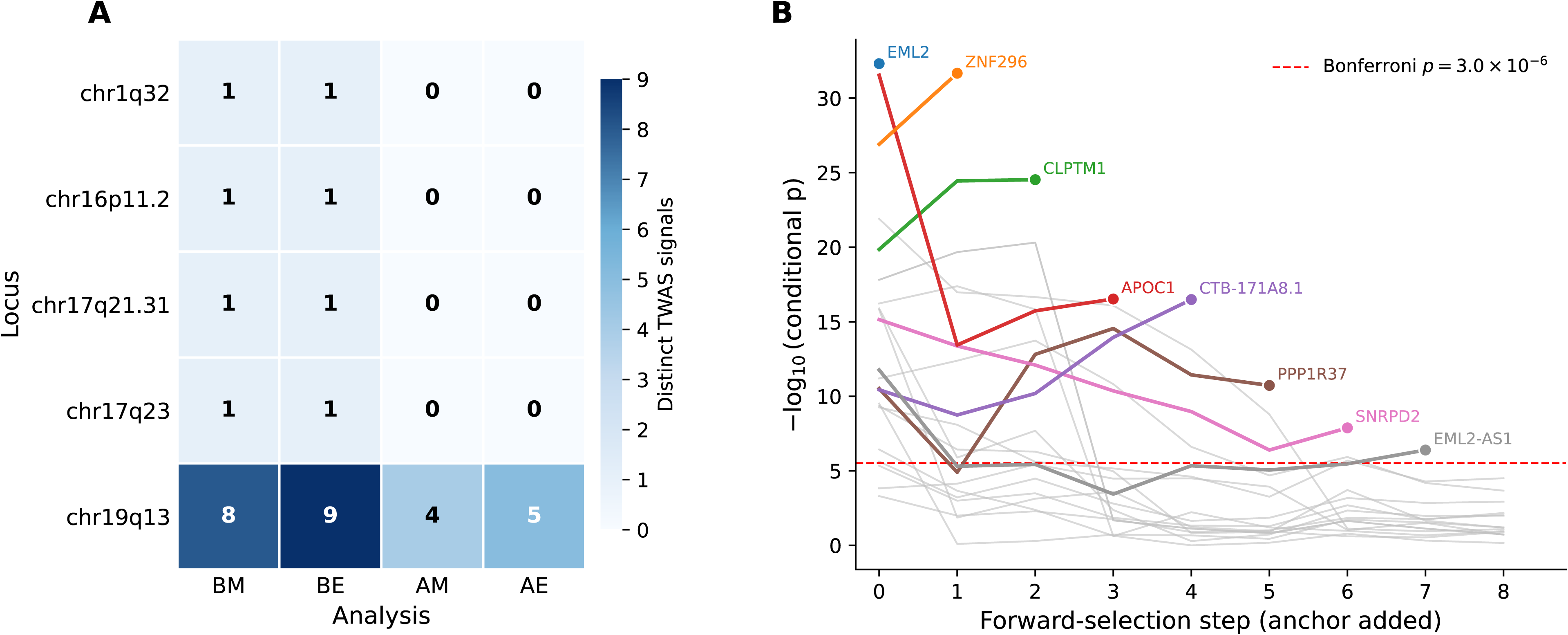
Comparison of association signals between the two retinal eQTL panels. Each point represents one of the 62 unique discovery genes, plotted by-log_lO_(p) from Bellenguez et al. + Meta-eQTL (x-axis) versus Bellenguez et al. + EyeGEx (y-axis). Dashed lines indicate Bonferroni significance thresholds for the Meta-eQTL and EyeGEx panels.

### Independent regulatory signals within the chromosome 19 cluster

Among the 62 discovery genes, 34 (55%) map to a dense cluster spanning approximately 1.7 Mb on chr19q13 (**Figure 3**). Since this region encompasses the *APOE* locus, we sought to determine whether these associations are independent. We first assessed pairwise correlations of predicted gene expression across this locus. Most gene pairs exhibited weak correlations, with only modest local structure among nearby genes **(Supplementary Figure 2)**. This suggested that the signals are not uniformly driven by a single shared expression component. To formally quantify the number of independent signals, we performed forward-selection conditional analysis (see **Methods**). At chromosome 19q13, we identified 8 conditionally independent signals in the Meta-eQTL discovery analyses (BM) and 9 in the EyeGEx analysis (BE), with 4 (for AM) and 5 (for AE) remaining in the validation cohort (**Figure 5A, Supplementary Table 9**). As an example, the sequential selection trajectory for the BM analysis showed distinct biological roles, including *ZNF226*, which regulates transcription, and *CLPTM1*, which has been prioritized in multiple AD GWAS and TWAS [34] (**Figure 5B**). In contrast, all tested multi-gene loci outside of chromosome 19q13 harbored a single conditionally independent signal in discovery (**Figure 5A**). Together, this functional diversity, combined with the persistence of multiple conditionally independent signals, supports the interpretation that the chr19q13 associations extend beyond gene expression correlation with *APOE* and implicate multiple partially independent regulatory mechanisms [43].

**Figure 5.**
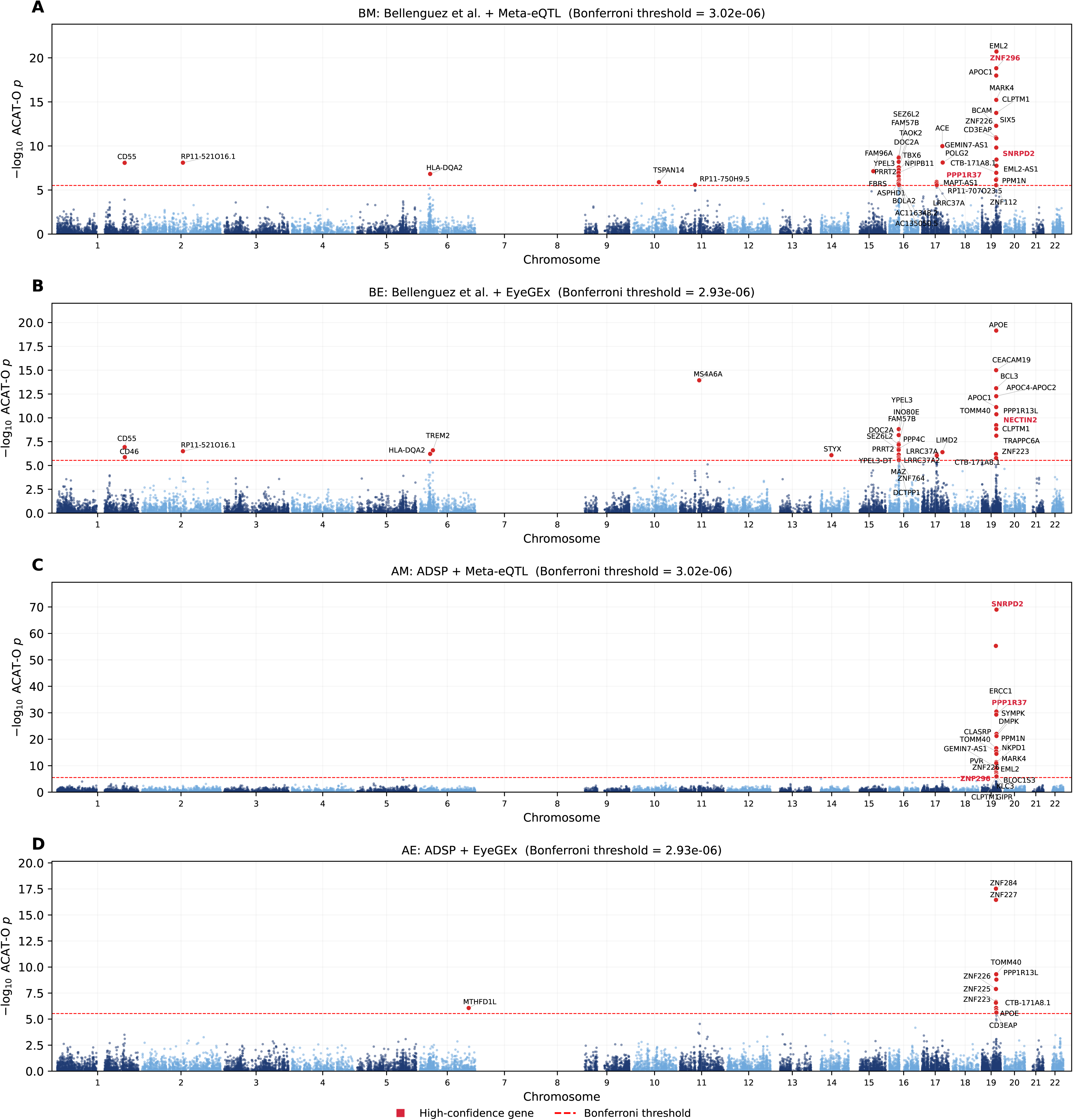
Forward-selection conditional analysis reveals polygenic architecture at chr19q13 and single distinct signals at four non-APOE loci. (A) Number of distinct TWAS signals per locus across the four analyses. (B) Conditional-log_lO_(p) across forward-selection steps at chr19q13 in Bellenguez et al. + Meta-eQTL. At each step, the top gene is selected as an anchor and removed, and remaining genes are re-tested conditional on the cumulative anchor set using the genetically predicted expression covariance matrix. The eight selected anchors (*EML2, ZNF296, CLPTM1, APOC1, CTB-171A8.1, PPP1R37, SNRPD2, EML2-AS1*) are colored in selection order; non-significant genes are shown in gray. The dashed red line marks the BM Bonferroni threshold (p = 3.0 × 10^-6^).

### Colocalization analyses identify shared causal variants

Finally, we conducted colocalization analysis to determine whether our significant genes share a single causal variant. Across discovery and validation analyses, we identified five unique genes (seven gene–eQTL panel pairs) from Bellenguez et al. discovery statistics that demonstrated strong colocalization (PP.H4 > 0.75), mapping to three loci on chromosomes 16, 17, and 19 (**Supplementary Figure 3, Supplementary Table 10**). No colocalizations were observed with ADSP, consistent with the smaller GWAS sample size reducing power for this analysis. The highest posterior probabilities were observed at chromosome 17q21.31 *MAPT* inversion region, where the paralogs *LRRC37A* and *LRRC37A2* shared a causal variant with AD risk in both eQTL panels (PP.H4 > 0.96; leading variants rs2532240 in Meta-eQTL and rs2696466 in EyeGEx; **Supplementary Figure 3A–B**). However, because the two genes share the same lead variants, colocalization alone cannot distinguish which paralog independently drives the association. Beyond some expected colocalizations, such as the canonical AD risk gene *ACE* in the Meta-eQTL panel (PP.H4 = 0.786, leading variant rs4292; **Supplementary Figure 3C**), the EyeGEx dataset captured two uniquely compelling causal signals at *APOE* locus (PP.H4 = 0.764, leading variant rs7247227; **Supplementary Figure 3D**) and *PPP4C* locus at chr16p11.2 (PP.H4 = 0.750, leading variant rs4238959; **Supplementary Figure 3E**).

## Discussion

In this study, we investigated the shared genetic architecture between the retina and the brain in AD by conducting a retinal TWAS. To the best of our knowledge, no other study has evaluated AD risk using retinal bulk tissue via TWAS yet. By integrating the largest available AD GWAS meta-analysis with two independent retinal eQTL panels, we identified 62 unique AD-associated genes. To identify high-confidence associations, we implemented a matched-panel validation strategy using the independent ADSP cohort, replicating 31 unique genes. Our findings demonstrate that genetically predicted expression in the retina captures both established AD risk loci and novel candidates. From a translational perspective, this molecular overlap highlights the retina’s potential as an accessible window into AD pathology, which is typically observable only in postmortem brain tissue.

Our results provide biological insights into the retina-brain neurodegenerative axis, most notably through the convergent identification of immune and complement regulatory mechanisms. We identified significant associations for *TREM2* and *MS4A6A*, genes centrally involved in microglial activation [44]. Their identification here aligns with the presence of resident myeloid cells in the retina, suggesting that they exhibit activation patterns analogous to those of brain microglia during neurodegenerative stress [44]. Furthermore, the robust cross-panel validation of *CD55* and *CD46* within regulators of the complement activation cluster on chromosome 1q32 highlights critical complement pathway dysregulation [45]. This supports growing evidence that complement activation contributes to synaptic loss via microglial pruning [45]. Because complement overactivation is a hallmark of both AD and AMD, these findings provide a direct transcriptomic link reinforcing the hypothesis of shared immune-mediated neuronal damage across aging neural tissues.

A known limitation of TWAS is that correlated predicted expression among nearby genes can produce co-significant associations that do not reflect independent causal effects [46]. This is particularly relevant at chr19q13, but our conditional analysis disentangled this region, identifying multiple conditionally independent signals across both discovery analyses and confirming that *CLPTM1*, *ZNF226*, and *CEACAM16-AS1* represent distinct regulatory effects extending beyond *APOE*. Furthermore, rather than only identifying association, we further extend our analysis to check the direction concordance of TWAS effect sizes. Our cross-cohort analysis confirmed that most of the genes (36 out of 39) were directionally concordant with only 3 exceptions. We further proved that 2 of them (*ZNF296* and *SNRPD2*) were statistical artifacts driven by extreme *APOE* genetic architecture and differential cohort power, with only *TOMM40* retaining genuine LD-driven discordance. We consider this likely reflects LD complexity at the *APOE/TOMM40* haplotype. In such dense genetic regions, the tight clustering of multiple regulatory variants can produce divergent, cohort-specific eQTL effect estimates across independent datasets. These observations align with growing recognition that TWAS effect directions are less stable than association p-values [47]. Such directional discordances are frequently driven by non-trait-relevant eQTLs tagging disease haplotypes through distinct, tightly linked regulatory variants [46,47]. Furthermore, these discrepancies can be heavily exacerbated by technical variations in the RNA-seq quantification methods and transcriptomic annotations used to build multi-SNP expression predictions [48].Genuine biological phenomena also play a role, as tissue-specific opposite directional eQTL effects have been documented even between closely related tissue sub-regions [49]. Consequently, effect directions in dense-LD settings warrant explicit sensitivity adjudication.

Beyond established loci, our analysis identified several novel and biologically notable candidates. For example, using EyeGEx, we identified *STYX*, a pseudophosphatase involved in neuronal growth. MR provided strong evidence that genetically predicted *STYX* expression is causally associated with increased AD risk, with no pleiotropic effect in discovery. Additionally, the Meta-eQTL panel highlighted foundational cellular maintenance mechanisms operating within the retina during AD pathogenesis, prioritizing novel genes linked to mitochondrial DNA replication (*POLG2*) and ribosomal biogenesis (*POLR1G*).

The complementary profiles of the two retinal eQTL panels, which showed 19% overlap despite profiling the same tissue type, underscore the value of integrating multiple eQTL references. The Meta-eQTL panel produced stronger signals at several loci on chromosome 19, while the EyeGEx panel uniquely captured canonical genes such as *APOE, MS4A6A,* and *TOMM40*. A plausible explanation is that EyeGEx includes AMD-affected retinas across four disease stages, which may unmask the regulatory variants that transcriptionally remain silent in the healthy tissue profiled by the Meta-eQTL panel [17]. Integration of multiple independent references is therefore critical for maximizing the capture of AD-relevant transcriptomic variation.

Despite these insights, several limitations of our analysis should be considered. First, both eQTL reference panels used bulk RNA sequencing. Since the retina is highly heterogeneous, future single-cell eQTL resources will be essential for identifying the precise cell populations driving these associations. Second, the phenotype definitions differ across the two AD GWAS datasets: Bellenguez et al. included proxy cases, while ADSP was restricted to clinically diagnosed cases. Third, for several novel findings from the Meta-eQTL panel (*POLR1G, POLG2, GEMIN7-AS1*, and ENSG00000279095), the genetic architecture of their expression regulation is polygenic, with no single genome-wide significant eVariant. While TWAS models aggregate these moderate signals to achieve predictive power [50], our application of MR with a relaxed instrument variable threshold (FDR < 0.1) increases the risk of bias and horizontal pleiotropy. Finally, our study was restricted to European ancestry, limiting generalizability to diverse populations.

In conclusion, our retinal TWAS demonstrates that expression-based analyses in retinal tissue capture both established and previously underdocumented components of AD genetic architecture. These findings motivate several future directions, including longitudinal retinal imaging studies to link molecular signals to structural changes observable in vivo, highlighting the retina as a valuable and accessible system for investigating AD pathogenesis.

## Data Availability

The Meta-eQTL (Strunz et al.) retinal eQTL summary statistics are openly available at http://www-huge.uni-regensburg.de/ PLOS. The EyeGEx (Ratnapriya et al.) retinal eQTL summary statistics are available through NEI Commons and GEO under accession GSE115828. The Bellenguez et al. 2022 AD GWAS summary statistics were obtained from the NHGRI-EBI GWAS Catalog under accession GCST90027158 (https://www.ebi.ac.uk/gwas/studies/GCST90027158). The ADSP GWAS summary statistics generated for this study are available through NIAGADS (https://dss.niagads.org/) under controlled access.The TWAS summary statistics generated in our study will be deposited in the UNC Dataverse (https://dataverse.unc.edu/), upon publication.

## Acknowledgement

This work involves secondary analyses of the ADSP data. We thank the study participants, the ADSP project, and the NIAGADS Data Sharing Service for providing the valuable data. We also thank the Bellenguez et al. 2022 GWAS meta-analysis, as well as the Strunz et al. and EyeGEx retinal eQTL studies. This study was partially supported by NIH grants R01AG079291, R01AG085581, and R01AG075884.

## Author contributions

Y.Z., Y.L., and A.R.V.R.H conceptualized the project; Y.Z., J.D., H.S., and R.S implemented the pipeline, conducted formal analysis; B.C. curated the data; Y.Z., J.D. wrote the manuscript; A.R.V.R.H., Y.L., and J.D. supervised the projects; All authors reviewed and approved the manuscript.

## Software and data availability

The Meta-eQTL (Strunz et al.) retinal eQTL summary statistics are openly available at http://www-huge.uni-regensburg.de/PLOS. The EyeGEx (Ratnapriya et al.) retinal eQTL summary statistics are available through NEI Commons and GEO under accession GSE115828. The Bellenguez et al. 2022 AD GWAS summary statistics were obtained from the NHGRI-EBI GWAS Catalog under accession GCST90027158 (https://www.ebi.ac.uk/gwas/studies/GCST90027158). The ADSP GWAS summary statistics generated for this study are available through NIAGADS (https://dss.niagads.org/) under controlled access. All TWAS analyses were performed using OTTERS (available at https://github.com/daiqile96/OTTERS) [19]. Quality control procedures utilized PLINK v1.9 [51]. ACAT p-value combination was implemented in R version 4.1.0 [52]. All code and TWAS summary statistics will be made available at https://github.com/clairenyz/retinal-TWAS-AD. The TWAS summary statistics generated in our study will be deposited in the UNC Dataverse (https://dataverse.unc.edu/), upon publication.

