## Supplementary Figures for "Retinal Transcriptome-Wide Association Study Identifies Novel Alzheimer’s Disease Risk Genes"

**Supplementary Materials**

**Supplementary Methods**

**AD GWAS summary statistics**

**Discovery phase:** We used the largest AD GWAS meta-analysis in individuals of European ancestry to date, derived by Bellenguez et al. [1]. This data included 111,326 cases and 677,663 controls , comprising both clinically diagnosed cases and proxy cases (i.e., individuals with parental history of AD or dementia). The substantially larger sample size provides greater power for gene discovery than earlier GWAS. GWAS summary statistics for 21,058,986 markers were downloaded for this study, with a median sample size of 471,522.

**Validation phase:** We validated our findings in the ADSP study, which comprises individual-level data on approximately 10,000 clinically diagnosed AD cases and 11,000 controls. We included only participants with non-missing AD status who self-identified as White, yielding a final sample of 6,730 cases and 5,381 controls. We restricted our analysis to variants with minor allele frequency (MAF) > 0.01 and genotype missingness < 0.95. For 991 individuals older than 90, we capped their age at 90, and for individuals with missing age data, we imputed with the mean age (73.9 years). We then conducted a GWAS for AD using REGENIE [2], adjusting for age, sex, and the top 10 principal components (PCs). Validated gene lists were derived by requiring nominal significance (p < 0.05) in the matched eQTL analysis (i.e., Bellenguez et al.+Meta-eQTL discoveries validated in ADSP+Meta-eQTL; Bellenguez et al.+EyeGEx discoveries validated in ADSP+EyeGEx), as well as consistent effect direction (details see below).

**Retinal eQTL panels**

**Meta-eQTL retinal eQTL panel:** This eQTL dataset represents a mega-analysis combining genotype and RNA-sequencing data from 311 healthy postmortem retinal samples of European ancestry across three independent study sites [3]. Briefly, raw reads were uniformly reprocessed, aligned to Ensembl genome build GRCh38, and normalized using trimmed mean of M-values (TMM), with ComBat applied to correct for study-site batch effects. Expression was measured from bulk retinal tissue comprising multiple cell types (photoreceptors, retinal ganglion cells, bipolar cells, Müller glia, retinal pigment epithelium, and microglia-like resident macrophages) without cell-type deconvolution. The *cis*-eQTL mapping (variants within $\pm$1 Mb of the gene transcription start site [TSS]) identified 403,151 significant eVariants regulating 3,007 eGenes at a false discovery rate (FDR) $<$ 0.05, with 744 additional independent secondary signals detected through conditional analysis. Since there is currently no single-cell eQTL data for human retinal tissue, this Meta-eQTL panel represents the largest available resource of healthy retina eQTL.

**EyeGEx retinal eQTL panel [4]:** This data was generated from 406 postmortem retinal samples of European ancestry donors, including both controls (MGS1, n = 105) and individuals at distinct stages of age-related macular degeneration (MGS2–4, n=348), graded according to the Minnesota Grading System (MGS). Same as the Meta-eQTL panel, expression was measured in bulk retinal tissue without cell-type resolution. RNA-sequencing yielded a median of 32.5 million uniquely mapped paired-end reads per sample. The *cis*-eQTL analysis, performed with adjustment for disease status, age, sex, population stratification, and batch effects, identified 14,565 eVariants controlling expression of 10,474 eGenes (8,529 protein-coding and 1,358 noncoding) at FDR $\leq$ 0.05, including 4,541 eQTLs detected only in the retina relative to Genotype-Tissue Expression (GTEx) tissues.

Both retinal eQTL reference panels were quality-controlled (QCed) by their original studies prior to public release. For the Meta-eQTL panel (n = 311 retinas), Strunz et al. applied genotype and RNA-seq QC, filtered variants at MAF > 0.01, and retained genes with count per million (CPM) > 1 in at least 10% of samples per constituent dataset, yielding 16,766 autosomal genes with eQTL summary statistics. For the EyeGEx panel (n = 406 retinas), Ratnapriya et al. applied analogous sample-level and variant-level QC (MAF > 0.05), retaining 13,662 protein-coding and 1,462 noncoding genes. Only autosomal genes with available *cis*-eQTL summary statistics from each panel were included in downstream TWAS analyses.

**Gene annotation**

Gene coordinates for OTTERS input were derived from the Ensembl genome build GRCh38, accessed via the R packages *AnnotationHub* and *ensembldb* [5,6]. Homo sapiens EnsDb records (assembly GRCh38; AnnotationHub resource AH119325 or the most recent available equivalent) were queried to obtain transcript start and end positions for all annotated genes. Only autosomal genes were retained. *Cis*-eQTL windows were defined as $\pm$1 Mb from the gene TSS. Per-chromosome annotation files were generated and provided as input to OTTERS, along with the corresponding eQTL and GWAS summary statistics.

**TWAS analysis using OTTERS**

We performed TWAS using the OTTERS [7] in a two-stage framework, where stage 1 leverages retinal eQTL summary statistics to train multiple expression-prediction models, and stage 2 integrates these models with AD GWAS data to evaluate gene expression-disease associations and aggregate results into a single statistical inference via an Aggregated Cauchy Association Test (ACAT-O) [8]. Separate TWAS analyses were conducted for each combination of GWAS datasets (Bellenguez et al., ADSP) and eQTL panels (Meta-eQTL, EyeGEx).

**Stage 1: eQTL Weight Estimation:** We estimated *cis*-eQTL weights using summary-level eQTL data (marginal effect sizes and p-values) from each retinal reference panel, along with the FUSION LDREF linkage disequilibrium (LD) reference panel, derived from the European-ancestry samples of the 1000 Genomes Project (1000G; n = 489) and lifted to GRCh38 [9]. For each gene, OTTERS trains expression imputation models using four methods, yielding five individual models: P+T [10] (P-value thresholding with LD clumping at two thresholds, p < 0.01 and p < 0.05, treated as two separate models), lassosum [11] (frequentist penalized regression), SDPR [12] (Bayesian nonparametric Dirichlet process regression), and PRS-CS [13] (Bayesian regression with continuous shrinkage priors).

**Stage 2: Gene-Trait Association Testing and Omnibus Testing:** For each of the five models, genetically regulated expression (GReX) was imputed into the GWAS cohorts using the estimated eQTL weights, and the resulting GReX was tested for association with AD risk using GWAS summary statistics. The FUSION Z-score statistic was used for summary-level GWAS input:

$$Z_{g}=\frac{\sum_{j=1}^{J} \hat{w}_{j}Z_{j}}{\sqrt{\hat{w}^{\top}V\hat{w}}}$$

where $\hat{w}_{j}$ is the estimated eQTL weight for SNP $j$, $Z_{j}$ is the single-variant GWAS Z-score for SNP $j$, $\hat{w}$ is the vector of eQTL weights across all $J$ overlapping SNPs, and $V$ is the genotype correlation (i.e., LD) matrix estimated from the 1000G reference panel.

In order to get a final p-value for each gene, OTTERS combines the five individual TWAS $p$-values using the ACAT-O [8]:

$$T_{ACAT-O}=\sum_{i=1}^{5} w_{i}tan\left[ \left( \frac{1}{2}-p_{i} \right)\pi\right]$$

where $p_{i}$ is the TWAS $p$-value from the $i$-th model, $w_{i}=1/5$ are equal weights assigned to each model, and $\pi$ is the mathematical constant. The resulting statistic follows an approximate Cauchy distribution under the null hypothesis, from which a final $p$-value is obtained.

To account for multiple testing, we applied Bonferroni corrections based on the number of genes successfully tested in each eQTL panel. Specifically, we tested 16,559 genes in the Meta-eQTL panel and 17,071 genes in the EyeGEx, yielding Bonferroni-corrected significance thresholds of 3.02 × 10^-6^ and 2.93 × 10^-6^, respectively (**Supplementary Table 1**). Finally, to provide a single and integrated set of summary statistics per gene per GWAS, we applied ACAT-O to the outputs of two retinal eQTL panels, yielding a combined value that accounts for potential correlations between the two panels.

### **Cross-cohort directional concordance analysis**

We assessed the consistency of the effect direction between the discovery and validation cohorts, by evaluating the sign of the mean FUSION Z-score, Z̄ (defined as the numerator $\hat{w}^{\top}Z_{GWAS}$ of FUSION Z-score formula, averaged across all five models). Since directional flips can arise from technical discrepancies, we evaluated any flipped genes using two specific criteria. First, we addressed the extreme statistical outliers at the *APOE* locus. The summary statistics from Bellenguez et al. were capped at |Z| = 9.3, whereas the ADSP summary statistics remained uncapped, reaching values up to 31.9 at the *APOE* index variant rs4420638. For genes whose eQTL weights load heavily on this region, this discrepancy disproportionately inflates the ADSP TWAS numerator (ŵᵀZ) and can artificially reverse the overall direction of effect. To correct this, we recalculated the validation Z̄ after constraining ADSP Z-scores to ±9.3 to match the discovery data, and quantified the fractional contribution of variants within ±700 kb of rs4420638 to flag loci driven by *APOE* LD (**Supplementary Table 7**). The ±700 kb window was chosen to cover the empirically observed LD extent of rs4420638 in the 1000G European-ancestry reference panel, within which r^2^ > 0.1 is sustained across the *APOE/TOMM40/APOC1* haplotype block. This window lies within the ±1 Mb *cis*-eQTL definition used elsewhere in our analysis and captures all HapMap3 variants likely to contribute meaningful LD leakage to TWAS weights for genes whose eQTL models load on the *APOE* region. Second, we accounted for differential statistical power between the discovery and validation cohorts using effective sample size, defined for case-control studies as N_eff_ *=* $\frac{4}{\frac{1}{N_{case}}+\frac{1}{N_{control}}}$. Effective sample sizes were N_eff_ ≈ 11,961 for ADSP (6,730 cases / 5,381 controls) and N_eff_ ≈ 382,472 for Bellenguez et al. (111,326 cases / 677,663 controls). Assuming a true shared effect, the validation Z̄ is expected to attenuate relative to the discovery Z̄ by a factor of $\sqrt{\frac{N_{ADSP}}{N_{Bellenguez et al.}}} \approx0.177$. We evaluated the residual between the observed validation Z̄ and the expected attenuated discovery Z̄. Finally, a discordance was classified as genuine biological heterogeneity only if the sign reversal persisted after Z-score capping and the residual deviated from zero by more than two standard errors. Otherwise, the directional flip was attributed to technical artifacts such as extreme-value distortion or differential statistical power.

**Pairwise correlations of predicted gene expression**

To assess the extent to which TWAS signals at multi-gene loci could be driven by shared predicted expression rather than independent regulatory effects, we computed pairwise Pearson correlations of GReX between all genes within each locus of interest. For each gene, GReX was calculated for each individual in the 1000 Genomes European-ancestry reference panel by multiplying the eQTL weights from the best-performing OTTERS model by individual-level genotype dosages at all model SNPs, then summing across SNPs. Pairwise Pearson correlation coefficients (r) were then computed across individuals between GReX vectors of all gene pairs at the locus. This procedure was applied separately to the Meta-eQTL and EyeGEx weight sets. Correlation matrices were ordered by chromosomal position and visualized as heatmaps.

**Conditional analyses within multi-gene loci**

To estimate the number of conditionally independent TWAS signals per locus, we performed forward-selection conditional TWAS using summary statistics. For each locus (i.e., a cluster of Bonferroni-significant genes with TSS within 1 Mb on the same chromosome), we computed the GReX-GReX covariance matrix $C = W^{\top}VW$, where $W$is the best-performing OTTERS model's eQTL weights for each gene and $V$ is the 1000G European ancestry LD matrix restricted to SNPs present in both the weight and LD files. The conditional Z-score for gene *j* given an anchor set *S* was computed as:

$$Z_{\left( j | S \right)}=\frac{\left( Z_{j}- R_{j,S}R_{S,S}^{-1}Z_{S} \right)}{\sqrt{1 - R_{j,S}R_{S,S}^{-1}R_{S,j}}}$$

where *Z_j_* is the marginal FUSION *Z*-score for gene *j*, Z*_S_* is the vector of marginal FUSION *Z*-scores for genes in the anchor set *S*. The gene-level correlation matrix, R, was obtained by standardizing the covariance matrix C, such that each element $R_{ij} = \frac{C_{ij}}{\sqrt{C_{ii}C_{jj}}}$. Within this framework, R*_j,S_* is the row vector capturing the correlation between gene *j* and the anchor genes, and R*_S,S_* is the correlation submatrix among the anchor genes.

We applied iterative forward selection: at each step, the gene with the smallest conditional p-value was added to *S*, and the procedure terminated when no remaining gene reached the panel-wide Bonferroni threshold. The final cardinality of *S* was reported as the number of distinct signals at that locus. The procedure was applied separately to each of the four analysis configurations. Five multi-gene loci were tested: chr1q32 (*CD55/CD46*; 207.32–207.80 Mb), chr16p11.2 (*SEZ6L2/DOC2A* region; 29.38–31.12 Mb), chr17q21.31 (*MAPT/LRRC37A* region; 45.60–46.56 Mb), chr17q23 (*ACE* region; 63.48–64.50 Mb), and chr19q13 (*APOE* region; 44.05–45.77 Mb).

**Supplementary Figures**

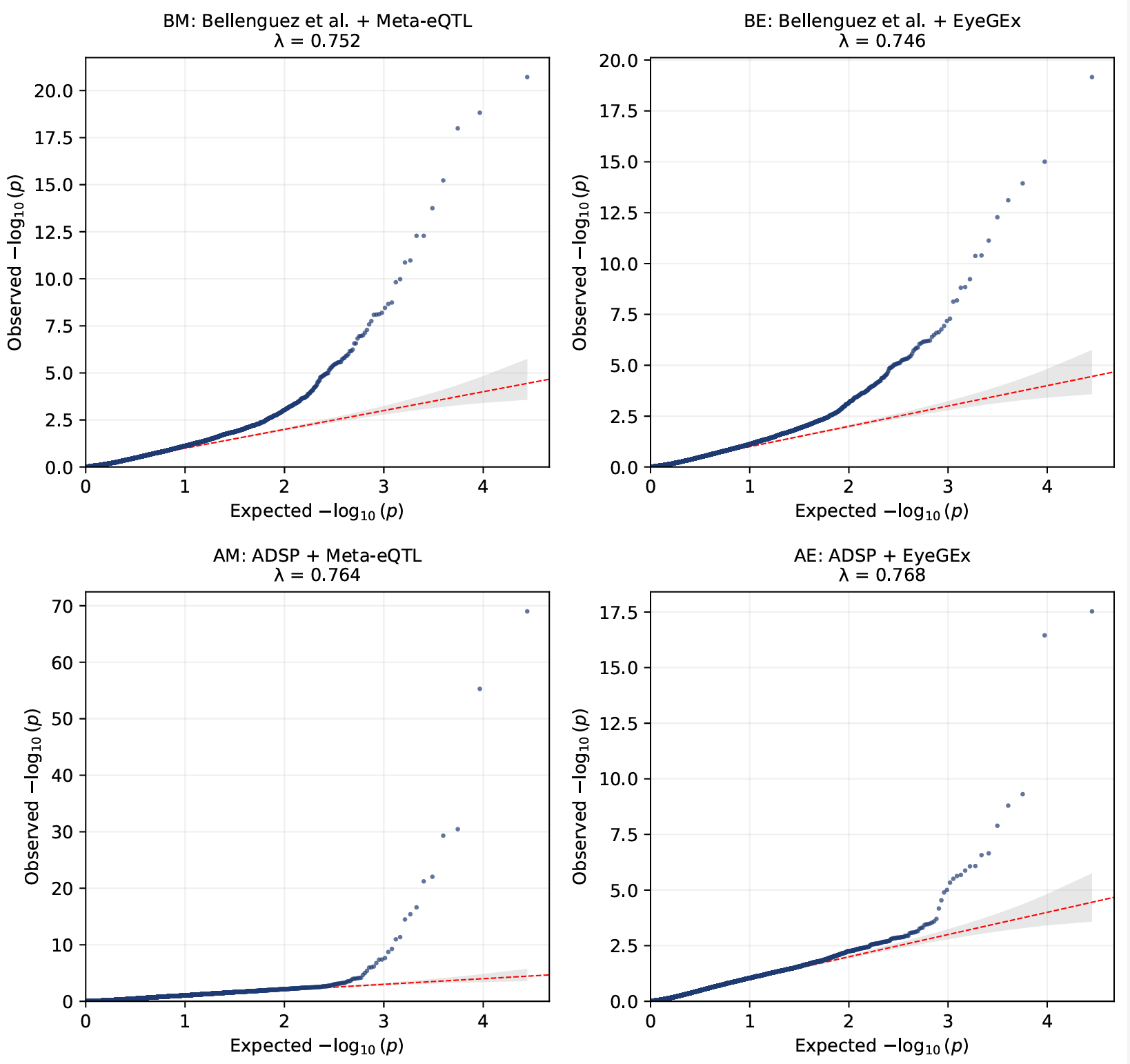
**Supplementary Figure 1. Quantile–quantile (QQ) plots of TWAS association statistics across four analyses.** Results are shown for BM, BE, AM, and AE analyses. Observed $-\log_{10}(p)$ values from OTTERS’ ACAT-O gene-level tests are plotted against expected values under the null hypothesis. Genomic inflation factors ($\lambda$) are shown in each panel. Overall deviation from the null distribution is modest (with consistent deflation observed) and primarily driven by loci with strong association signals.

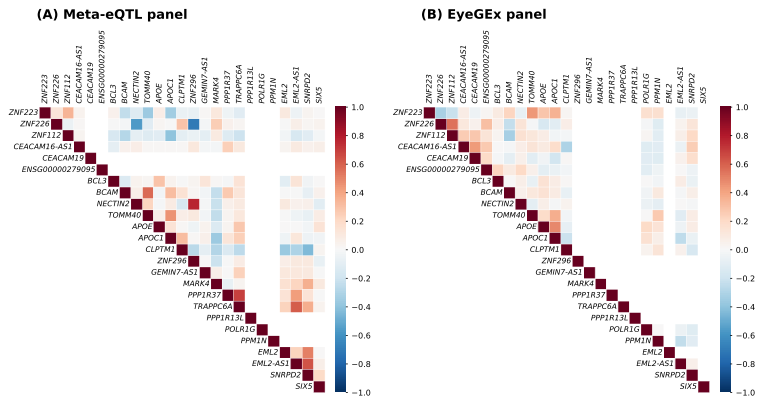

**Supplementary Figure 2. Pairwise correlation of genetically regulated expression (GReX) among nearby genes on chromosome 19 locus.** Pearson correlation matrices were computed using OTTERS predicted expression (GReX) for chromosome 19 locus genes (44–55 Mb, GRCh38), ordered by physical position. Panels show the Meta-eQTL (A) and EyeGEx (B) eQTL-weight models. Color indicates Pearson $r$ from $-1$ (blue) to $+1$ (red).

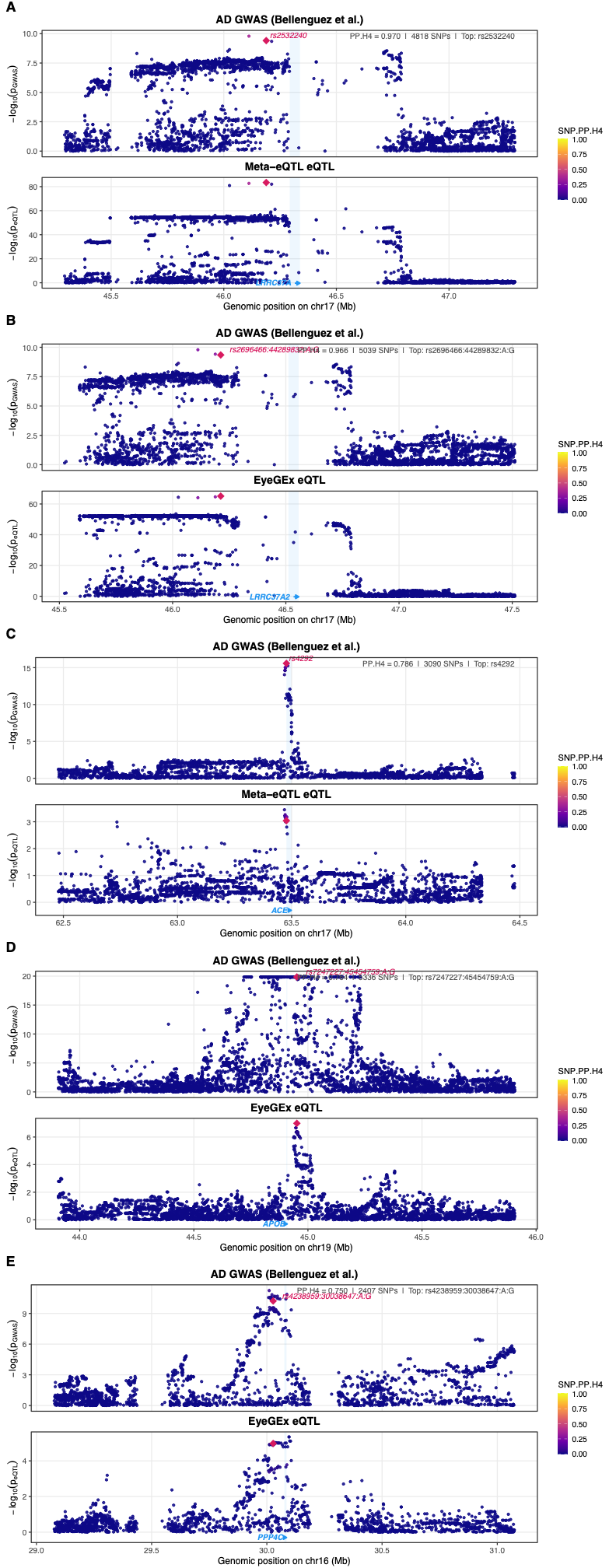

**Supplementary Figure 3. Colocalization of retinal eQTL and AD GWAS signals.** Regional association plots showing AD GWAS −log₁₀(p) (top) and retinal eQTL −log₁₀(p) (bottom). Purple diamonds indicate the lead colocalizing variant. **(A)** *LRRC37A* at 17q21.31, Bellenguez et al. + Meta-eQTL (PP.H4 = 0.970, rs2532240). **(B)** *LRRC37A* at 17q21.31, Bellenguez et al. + EyeGEx (PP.H4 = 0.968, rs2696466; r^2^ = 0.997 with rs2532240). **(C)** *ACE* at 17q23.3, Bellenguez et al. + Meta-eQTL (PP.H4 = 0.786, rs4292). **(D)** *APOE* at 19q13.32, Bellenguez et al. + EyeGEx (PP.H4 = 0.764, rs7247227). **(E)** *PPP4C* at 16p11.2, Bellenguez et al. + EyeGEx (PP.H4 = 0.750, rs4238959).
